# Latest Generation Colonoscope Yields Improvements in Cecal Insertion Time and Adenoma Detection Rate: A Retrospective Comparison of Two Colonoscope Brands

**DOI:** 10.1101/2021.05.01.21256464

**Authors:** Pauline Yasmeh, Joseph J. Vicari, Aaron J. Shiels

## Abstract

**Background and Aims:** High quality colonoscopy remains the cornerstone of colon cancer prevention. Studies have shown that generational advances in colonoscopes result in more favorable clinical outcomes. Performance of various endoscopes is determined using objective quality measures. The aim of this study was to compare these measures between two colonoscope platforms.

**Methods:** This study is a single center retrospective study of 3,761 patients undergoing initial screening colonoscopy between November 2013 and May 2020 using two different colonoscope platforms (Fujifilm EC-760R-V/L, n=2287 and Olympus CF and PCF 180 series, n= 1474). The primary outcomes measured were cecal insertion time, withdrawal time, and adenoma detection rate.

**Results:** Procedures completed with the Fujifilm colonoscope had mean cecal insertion times that were 2.01 minutes shorter than procedures completed with Olympus (p<.0001). Procedures completed with Olympus brand colonoscopes had prolonged cecal insertion times (≥10 minutes) nearly three times as often compared to those performed with Fujifilm colonoscopes (p<.0001). Cecal withdrawal times were slightly longer for females performed with Fujifilm colonoscopes, but there was no difference when all procedures were included (p=.09). Procedures completed with Fujifilm colonoscopes had higher adenoma detection rates compared to those completed with Olympus colonoscopes (p<0.001).

**Conclusions:** In this study, Fujifilm colonoscopes outperformed Olympus colonoscopes in screening colonoscopies with statistically and clinically significant shorter cecal insertion times and higher adenoma detection, though both platforms had similar withdrawal times.

## Introduction

High quality colonoscopy remains the cornerstone of colon cancer prevention. Advances in colonoscopy have resulted in improvements in adenoma detection rate (ADR) and decreases in risk of colon cancer^1-2^. Improved bowel preparation, longer withdrawal times, and more detailed mucosal examination techniques have all contributed to higher quality colonoscopy^3-5^.

The typical endoscopist spends considerable time and effort performing colonoscopy. Even small advances in endoscopist performance can translate into significant improvements in outcomes given the number of colonoscopies performed worldwide. Studies have shown that generational advances in colonoscopes from the same manufacturer result in more favorable clinical outcomes, including adenoma detection rates^6-8^.

However, there are limited studies comparing the performance of various colonoscopy platforms. A single center prospective study of 3000 colonoscopies comparing Olympus and Pentax colonoscopes revealed no significant difference in cecal intubation rates, withdrawal times, or adenoma detection rates^9^. Another smaller study with a single endoscopist also failed to demonstrate a difference between Olympus and Pentax colonoscopes^10^.

Colonoscope performance from the endoscopist’s perspective is partially a subjective experience which makes comparison challenging. The ease and speed of cecal insertion time provides one objective measure by which colonoscopes can be compared. Decreases in cecal insertion time can translate into lower overall procedure times and greater efficiency in the endoscopy facility. Longer cecal insertion time may also be associated with decreased adenoma detection rate^11^. We sought to compare two colonoscopy platforms (Fujinon, Fujifilm, Tokyo, Japan and Olympus, Tokyo, Japan) to determine if clinically relevant differences could be identified.

## Methods

A single center retrospective study was completed. Approval was obtained from the Institutional Review Board at the University of Illinois-College of Medicine at Rockford. All colonoscopies were performed at Rockford Gastroenterology Associates (RGA) in the ambulatory surgical center between November 2013 and May 2020. The procedures were performed by the same 11 gastroenterologists. All endoscopists had at least 3 years of experience in practice and had performed more than 3000 colonoscopies prior to the study.

All patients were 50-75 years of age, undergoing their initial screening colonoscopy. There were no restrictions on the type of bowel preparation utilized. Sedation and analgesia were accomplished most commonly with moderate sedation using fentanyl and midazolam. Monitored anesthesia care for deep sedation with propofol was less frequently utilized. Patients were excluded from the analysis for inadequate bowel prep, incomplete colonoscopy, or incomplete data on insertion or withdrawal times.

Olympus colonoscopes (CF 180 series, PCF 180 series) were exclusively used from November 2013 to November 2018. The choice of colonoscope was at the discretion of the endoscopist. The entire endoscopy facility converted to Fujifilm colonoscopes (EC-760R-V/L) in November 2018 and data was collected through May 2020. To minimize the effects of colonoscope wear, only up to the first two years of each colonoscope’s lifetime were included in the analysis. Refurbished or damaged colonoscopes were excluded.

Procedural data was recorded by the endoscopist and nurse through the NextGen EHR system and automatically uploaded into GiQuic. This database was then queried for the desired information and all data was deidentified for analysis. Demographics were recorded for each patient, including age, sex, and body mass index (BMI). Cecal insertion time was automatically calculated by the EHR as the time from rectal insertion to cecal intubation. Withdrawal time was time from cecal intubation to removal of the colonoscope from the patient.

The primary outcomes were cecal insertion time, withdrawal time, and adenoma detection rate. In addition to calculating mean cecal insertion time, the proportion of procedures with a cecal insertion time of at least 10 minutes was assessed. Analysis was performed by comparing Fujifilm and Olympus colonoscopes. Subgroup analysis was performed by sex and type of Olympus colonoscope (CF, PCF). Statistical analysis was performed using Student t-test for continuous variables and Chi-square test for categorical values. All authors had access to study data and participated in review of the manuscript.

## Results

### Baseline characteristics

Table 1 summarizes the baseline characteristics of the 3,761 patients included in the analysis. 1,474 patients underwent colonoscopy with an Olympus colonoscope. The adult colonoscope (CF) was utilized in 599 cases of which 435 (72.6%) were male. The pediatric colonoscope (PCF) was utilized in 875 cases of which 222 were male (25.4%). 2,287 patients underwent colonoscopy with a Fujifilm colonoscope (EC-760R-V/L), of which 1,157 (50.6%) were male. All groups were similar with regard to age and body mass index.

**Table 1.** Patient baseline characteristics.

| Table 1. Patient baseline characteristics |  |  |  |  |  |  |  |
| --- | --- | --- | --- | --- | --- | --- | --- |
|  | Fujifilm<br>(n=2287) | All Olympus<br>(n=1474) | p value | Olympus CF<br>(n=599) | p value | Olympus PCF<br>(n=875) | p value |
| Age | 55.6 (5.6) | 56.3 (6.0) | .0006 | 55.3 (5.3) | .2 | 57.0 (6.4) | <.0001 |
| BMI | 30.2 (6.7) | 29.6 (6.2) | .008 | 30.9 (6.0) | .02 | 28.7 (6.2) | <.0001 |
| Sex, M:F | 1157:1130 | 657:817 | .004 | 435:164 | <.0001 | 222:653 | <.0001 |

Mean (SD); p values represent comparison to Fujifilm

### Colonoscope specifications

Table 2 reviews the specifications of the various colonoscopes used in the study. The Fujifilm and Olympus colonoscopes are high definition with similar observation range, working length, variable stiffness capability, and tip deflection. The diameter of the Fujifilm colonoscope (11.8 mm) is intermediate between the Olympus CF (12.8 mm) and PCF colonoscopes (11.0 mm). The field of view is 170° for the Fujifilm and Olympus CF colonoscope and 140° for the Olympus PCF colonoscope.

**Table 2.** Colonoscope specifications.

| Table 2. Colonoscopy specifications |  |  |  |
| --- | --- | --- | --- |
|  | Fujifilm EC-760R-V/L | Olympus CF 180 series | Olympus PCF 180 series |
| Television standard | HD | HD | HD |
| Field of view | 170° | 170° | 140° |
| Observation range | 2-100 mm | 3-100 mm | 3-100 mm |
| Working length | 169 cm | 168 cm | 168 cm |
| Outer diameter | 12.0 mm | 12.8 mm | 11.8 mm |
| Channel diameter | 3.8 mm | 3.7 mm | 3.2 mm |
| Variable stiffness | Yes | Yes | Yes |
| Tip deflection | Up/down 180°, left/right 160° | Up/down 180°, left/right 160° | Up/down 180°, left/right 160° |

### Cecal insertion time

Procedures completed with the Fujifilm colonoscope had significantly lower cecal insertion times compared to procedures completed with Olympus colonoscopes overall (table 3). The cecal insertion time was 2.01 minutes less for Fujifilm (5.06 minutes, [95% CI: 4.84-5.18]) compared to Olympus (7.07 minutes, [95% CI: 6.82-7.32]) when all patients and colonoscope types were included (p<.0001). There was a 1.07 minute decrease in cecal insertion time when Fujifilm was compared to Olympus CF colonoscope (6.13 minutes, [95% CI: 5.77-6.49], p<.0001) and a 2.65 minute decrease when compared to Olympus PCF colonoscope (7.71 minutes, [95% CI: 7.38-8.04], p<.0001).

**Table 3.** Cecal insertion time.

| Table 3. Cecal insertion time |  |  |  |  |  |
| --- | --- | --- | --- | --- | --- |
|  | Fujifilm | All Olympus | Olympus CF | Olympus PCF | p value |
| All patients | 5.06 (4.84-5.18) | 7.07 (6.82-7.32) | 6.13 (5.77-6.49) | 7.71 (7.38-8.04) | <.0001 |
| Male | 4.48 (4.33-4.63) | 6.07 (5.72-6.42) | 5.61 (5.24-5.99) | 6.95 (6.27-7.63) | <.0001 |
| Female | 5.64 (5.45-5.83) | 7.87 (7.53-8.21) | 7.48 (6.66-8.30) | 7.97 (7.60-8.34) | <.0001 |

Mean time in minutes (95% CI); p value <.0001 for all 3 Olympus vs Fujifilm comparisons

In male patients, the cecal insertion time was 1.59 minutes less for procedures completed with Fujifilm (4.48 minutes, [95% CI: 4.33-4.63]) compared to those with Olympus overall (6.07, [95% CI: 5.72-6.42], p<.0001). There was a 1.13 minute decrease when Fujifilm was compared to Olympus CF colonoscope (5.61 minutes, [95% CI: 5.24-5.99], p<.0001) and a 2.47 minute decrease when compared to Olympus PCF colonoscope (6.95 minutes, [95% CI: 6.27-7.63], p<.0001).

In female patients, the cecal insertion time was 2.23 minutes less for procedures completed with Fujifilm (5.64 minutes, [95% CI: 5.45-5.83]) compared to those with Olympus overall (7.87 minutes, [95% CI: 7.53-8.21], p<.0001). There was a 1.84 minute decrease when Fujifilm was compared to Olympus CF colonoscope (7.48 minutes, [95% CI: 6.66-8.30], p<.0001) and a 2.33 minute decrease when compared to Olympus PCF colonoscope (7.97 minutes, [95% CI: 7.60-8.34], p<.0001).

Prolonged cecal insertion times (≥10 minutes) were significantly more frequent for procedures performed with Olympus colonoscopes compared to those performed with Fujifilm (table 4). This occurred in 18.3% [95% CI: 16.3-20.3] of procedures with an Olympus model colonoscope compared to 6.4% [95% CI:5.4-7.4] with a Fujifilm colonoscope (p<.0001). In males, this occurred in 12.2% [95% CI: 9.7-14.7] of Olympus cases and 4.2% [95% CI: 3.0-5.4] of Fujifilm cases (p<.0001). In females, there was a cecal insertion time of at least 10 minutes in 23.3% [95% CI: 20.4-26.2] of Olympus cases and 8.7% [95% CI:7.1-10.3] of Fujifilm cases (p<.0001).

**Table 4.** Cases with cecal insertion time ≥ 10 minutes.

| Table 4. Cases with cecal insertion time $\geq 10$ minutes | | | |
| --- | --- | --- | --- |
|  | Fujifilm | Olympus | p value |
| All patients | 6.4% (5.4-7.4) (147/2287) | 18.3% (16.3-20.3) (270/1374) | <.0001 |
| Male | 4.2% (3.0-5.4) (49/1157) | 12.2% (9.7-14.7) (80/657) | <.0001 |
| Female | 8.7% (7.1-10.3) (98/1130) | 23.3% (20.4-26.2) (190/817) | <.0001 |

Proportion (95% CI)

### Withdrawal time

There was no difference in withdrawal time when all colonoscopies performed with Fujifilm (12.4 minutes, 95% CI 12.2-12.6) were compared to those performed with Olympus (12.2 minutes, 95% CI 12.0-12.4, p=.09, table 5). For male patients, there was no difference in withdrawal time for procedures completed with Fujifilm (12.9 minutes, [95% CI: 12.6-13.2]) compared to those with Olympus (13.2 minutes, [95% CI: 12.8-13.7], p=0.23). For female patients, there was a slightly longer withdrawal time for Fujifilm procedures (12.0 minutes, [95% CI: 11.7-12.2]) compared to those with Olympus (11.3 minutes, [95% CI: 11.0-11.6], p=.0008).

**Table 5.** Withdrawal time.

| Table 5. Withdrawal time |  |  |  |
| --- | --- | --- | --- |
|  | Fujifilm | Olympus | p value |
| All patients | 12.4 (12.2-12.6) | 12.2 (12.0-12.4) | .09 |
| Male | 12.9 (12.6-13.2) | 13.2 (12.8-13.7) | .23 |
| Female | 12.0 (11.7-12.2) | 11.3 (11.0-11.6) | .0008 |

Mean time in minutes (95% CI)

### Adenoma detection rate

Colonoscopies performed with Fujifilm colonoscopes had a higher ADR compared to those performed with Olympus colonoscopes (table 6). This was statistically significant when all patients were included (Fujifilm 51.1% [95% CI: 49.1-53.1], Olympus 45.9% [95% CI: 43.4-48.4], p<.001). There was a trend toward higher ADR in males (Fujifilm 58.2% [95% CI: 55.4-61.0], Olympus 55.9% [95% CI: 52.1-59.7]) though this did not reach statistical significance (p=0.37). There was a statistically significant difference in ADR for females (Fujifilm 43.9% [95% CI: 41.0-46.8], Olympus 37.9% [95% CI:34.6-41.2], p=.01).

**Table 6.** Adenoma detection rate.

| Table 6. Adenoma detection rate |  |  |  |
| --- | --- | --- | --- |
|  | Fujifilm | Olympus | p value |
| All patients | 51.1% (49.1-53.1) (1169/2287) | 45.9% (43.4-48.4) (677/1474) | <.001 |
| Male | 58.2% (55.4-61.0) (673/1157) | 55.9% (52.1-59.7) (367/657) | .37 |
| Female | 43.9% (41.0-46.8) (496/1130) | 37.9% (34.6-41.2) (310/817) | .0098 |

Proportion (95% CI)

## Discussion

In this study, the Fujifilm EC-760R-V/L colonoscope outperformed the Olympus CF 180 series and PCF 180 series colonoscopes with regard to cecal insertion time and adenoma detection rate. There was, on average, a two minute decrease in the cecal insertion time and approximately a 5% increase in ADR. There was also a nearly three-fold increase in the number of cases with a prolonged cecal insertion time (≥10 minutes) for procedures performed with an Olympus colonoscope.

The reasons for the statistically and clinically significant different cecal insertion times appear to be related to intrinsic differences with the two brands of colonoscopes that cannot be gleaned from the published technical specifications. All study colonoscopes are high definition, variable stiffness, and have nearly identical working lengths and tip deflection. The field of view is 170° for the Fujifilm and Olympus CF and 140° for the Olympus PCF. To minimize the effects of colonoscope wear, only colonoscopes with up to two years of use were included and refurbished or damaged colonoscopes were excluded. The same 11 experienced endoscopists performed all study colonoscopies, eliminating endoscopist technique as a differentiating factor. All patients presented for their first screening colonoscopy and had similar BMIs. Similar types of preparation and sedation and analgesia were used throughout the study.

Improvements in ADR are associated with a decreased risk of colon cancer. In this study, the ADR was 5% higher for the Fujifilm colonoscope. A previous study suggested that longer cecal insertion times are associated with a decreased ADR^11^. Our findings support that hypothesis. The improvement in ADR was most pronounced in female patients undergoing colonoscopy with Fujifilm. This was also the group with the greatest difference in cecal insertion times.

Shorter insertion time allows more time for the withdrawal phase, where the vast majority of adenomas are detected and removed. There may also be some degree of endoscopist fatigue with longer cecal insertion time that can affect the ability to detect polyps during the withdrawal phase.

To date, there are limited studies comparing colonoscope performance between manufacturers. Given the subjective nature of colonoscope performance, there are limited objective measures for quantifying differences between colonoscopes. An experienced endoscopist can “feel” that an endoscope is a high quality instrument, but how can that be quantified, and how much difference does it make?

This study was prompted by the subjective experience between the RGA endoscopists and endoscopy nurses that the Fujifilm colonoscope outperformed the Olympus colonoscopes. This difference was noted soon after our endoscopy center changed to Fujifilm colonoscopes in November 2018. The endoscopists noted smoother, easier insertion of the colonoscope to the cecum, and our endoscopy nurses noted less need for abdominal pressure. Our goal was to test the hypothesis that the latest generation Fujifilm colonoscope was superior through objective measures.

Shorter cecal insertion time is an easily measured indicator of colonoscope performance that likely relates to smoother turns and less looping. There are advantages to a shorter cecal insertion time. First, there may be less patient discomfort as the insertion phase tends to be the most uncomfortable. Second, reducing time from the insertion phase allows additional time for the withdrawal phase, when most polyp detection is performed. Third, the endoscopy lab may run more efficiently by having decreased total procedure times. Although two minutes per case seems like a relatively short time, this becomes quickly noticeable throughout the course of a day at any high-volume facility. Also, decreasing the number of procedures with prolonged cecal insertion time improves the endoscopist’s efficiency in navigating a busy procedure schedule.

There are potential limitations to this study. Although the same 11 endoscopists performed all colonoscopies, it is reasonable to consider that their technique improved over 8 years. However, this would not seem to adequately explain the significant differences identified. We recognize that comparing the Fujifilm EC-760R-V/L to Olympus 180 series colonoscopes and not to the 190 series is a legitimate criticism. However, these findings should be encouraging to all endoscopists, as it suggests that colonoscope technology is making clinically meaningful advances.

This study provides the first comparison showing that one brand of colonoscope outperforms another. The goal of every endoscopist is to identify and remove adenomas and lower the risk of colon cancer through use of the best colonoscope technology. Future advances in colonoscopes should continue to focus on enhanced detection of colonic lesions, including adenomas. We believe that technological improvements can be best achieved through increased competition between colonoscope manufacturers. A large, prospective, multicenter head-to-head study between colonoscope manufacturers should be a top priority.

## Data Availability

All data referred to in the manuscript is saved and available.

## Acknowledgements

The authors thank Jason Pryor, Alana Hernandez, and Erica Natal for assistance in data acquisition. No external support or funding was received for this study.

## Notes

### Competing Interest Statement

The authors have declared no competing interest.

### Funding Statement

No external funding was received for this study.

### Author Declarations

Approval was obtained from the Institutional Review Board at the University of Illinois College of Medicine at Rockford.

